# Community transmission of rotavirus infection in a vaccinated population in Malawi: a prospective household cohort study

**DOI:** 10.1101/2020.04.05.20036574

**Authors:** Aisleen Bennett, Louisa Pollock, Naor Bar-Zeev, Joseph A. Lewnard, Khuzwayo C. Jere, Benjamin Lopman, Miren Iturriza-Gomara, Virginia E. Pitzer, Nigel A. Cunliffe

**Author notes:** **Corresponding author details:** Institute of Infection and Global Health, The Ronald Ross Building, University of Liverpool, 8 West Derby Street, Liverpool. Contributed equally (Virginia E. Pitzer, Nigel A. Cunliffe).

## Abstract

**Background:** Rotavirus vaccine effectiveness (VE) is reduced among children in low-income countries (LICs). Indirect (transmission-mediated) effects of rotavirus vaccine may contribute to the total population impact of vaccination. We estimated the effectiveness of rotavirus vaccine in preventing transmission of rotavirus to household contacts in Blantyre, Malawi.

**Methods:** We recruited vaccine-age-eligible children with acute rotavirus gastroenteritis (case-children), together with their household contacts. Clinical data and stool samples were collected from case-children at presentation, and prospectively from household contacts over 14 days. A single stool sample was collected from control households containing asymptomatic children age-matched to case-children. Samples were tested for rotavirus using real-time PCR. Risk factors for household transmission of rotavirus infection and clinical rotavirus disease were identified using logistic regression. Vaccine effectiveness against transmission (VE_T_) was estimated as one minus the ratio of secondary attack rates (SAR) in vaccinated and unvaccinated populations, using VE estimates from the associated diarrhoeal surveillance platform to estimate the counterfactual SAR without vaccination.

**Findings:** A total of 196 case-households and 55 control-households were recruited. Household SAR for rotavirus infection was high (65%); SAR for clinical disease was much lower (5%). Asymptomatic infection in control households was common (28%). Increasing disease severity was associated with increased risk of transmission of both rotavirus infection and disease to household contacts. Estimated VE_T_ was 39% (95% confidence interval 16-57%).

**Interpretation:** Rotavirus vaccine has the potential to substantially reduce household rotavirus transmission. This should be considered in clinical and health economic assessments of vaccine impact.

**Funding:** Wellcome Trust and NIH/NIAID.

## Introduction

Rotavirus vaccine has been introduced into over 90 countries worldwide, including 45 low- and middle-income countries (LMICs)^1^. However, rotavirus vaccine effectiveness (VE) is reduced in low-income countries (LICs), where disease burden is highest, compared to high-income countries^2^. Thus, despite high vaccine coverage, rotavirus remains the commonest cause of hospitalised diarrhoeal disease in some LICs^3^. Since direct VE is reduced in LMICs, additional transmission-mediated (indirect) effects of the vaccine have the potential to make important contributions to population-level vaccine impact. However, both the presence and extent of rotavirus indirect effects, and the mechanisms which underpin them, are poorly understood^4^.

Rotavirus vaccination mimics immunity induced by natural rotavirus infections, which confer incremental protection against severe rotavirus disease to a degree which varies by location^5–7^. Rotavirus disease severity has been demonstrated to correlate with faecal rotavirus shedding density in India and Malawi^8^, and available evidence suggests that severity of symptoms is related to the risk of transmission^9^. It is therefore plausible that vaccination, while not providing complete protection against disease, may reduce the severity of gastroenteritis and diminish viral shedding following exposure to natural rotavirus infection, leading to reduced infectiousness of an index case, a lower secondary attack rate (SAR) in exposed households, and reduced rotavirus transmission.

There are currently no published data describing household transmission of rotavirus from sub-Saharan Africa. Previous studies in high-income settings have demonstrated a high SAR within families and have highlighted the role of infants in introducing rotavirus infection into households^10,11^. A household transmission study in Ecuador demonstrated a SAR of 55% for asymptomatic infection^9^. However, extrapolation of data on rotavirus transmission from high- and middle-income countries to LICs is not appropriate because of fundamental differences in factors that may have a major influence on the risk of transmission such as living environments, crowding, contact patterns, access to sanitation systems, host immunity and frequency of exposure to rotavirus^4^.

This study aimed to investigate household rotavirus transmission in a semi-urban setting in Blantyre, Malawi, where the monovalent, G1P[8]-containing rotavirus vaccine (RV1), was introduced into the childhood immunisation programme in 2012. Our objectives were to (i) explore risk factors for transmission of rotavirus to household contacts, including symptom severity and faecal rotavirus shedding density, and (ii) estimate the effectiveness of rotavirus vaccine in preventing transmission of rotavirus infection to household contacts.

## Methods

### Recruitment and study design

We conducted a prospective cohort study across four government health facilities in Blantyre, Malawi between February 2015 and November 2016. These health facilities included Queen Elizabeth Central Hospital (QECH), the primary referral centre for Southern Malawi; and three health centres (Zingwangwa, Gateway, and Madziabango). Vaccine-age-eligible children presenting with acute gastroenteritis were screened for rotavirus using a point-of-care immuno-chromatographic rapid test (Rota-Strip, Coris BioConcept, Gembloux, Belgium). Children testing positive for rotavirus (case children) were recruited together with their household contacts, following informed consent.

In order to estimate SARs, households were followed prospectively for 14 days after symptom onset in the case child; this comprised active surveillance for clinical disease and stool collection from household contacts to detect rotavirus infection (case cohort). Control households were recruited in order to define the background prevalence of rotavirus infection in households in the community. Control households each contained an asymptomatic child frequency-matched based on age to case children, and were recruited from randomly generated Global Positioning System (GPS) locations in Blantyre district. Control households were excluded if there was a history of symptoms of gastroenteritis in any household member in two weeks prior to recruitment. Sample size estimates are detailed in the supplementary materials.

### Data collection

Clinical data were collected from case children including anthropometric measurements and assessment of disease severity using the standardised 20-point Vesikari score^12^. Demographic, risk factor and symptom questionnaires were completed for household contacts and control households upon recruitment. Receipt of rotavirus vaccine and HIV status were documented from government-issued, hand-held health passports.

Participants were defined as HIV infected if positive on HIV rapid test (children >12 months), or HIV DNA PCR (infants <12 months)^13^. Severe acute malnutrition (SAM) was defined using WHO criteria of weight-for-height Z score (WHZ) ≤3 standard deviations from the median or mid-upper arm circumference (MUAC) ≤11·5cm^14^.

### Sample collection

Case children with acute gastroenteritis had bulk stool and 1-2ml of serum collected at presentation for measurement of rotavirus viral load and anti-rotavirus immunoglobin A (IgA) titres, respectively. Stool samples were collected from household contacts at days 5-7 and 10-12 after symptom onset in the case child. Control household members had a single stool sample collected at the point of recruitment.

### Laboratory analysis

Faecal samples were tested for rotavirus using a VP6 semi-quantitative real-time PCR (qRT-PCR)^15^. A standard curve was included in each run to allow estimation of rotavirus viral load (copy numbers). Samples with Ct value >35 and <40 on VP6 PCR underwent confirmatory testing with a second qRT-PCR assay targeting the NSP3 gene^16^. Samples were defined as rotavirus positive if they contained ≥100 viral copy numbers and were positive on NSP3 assay. All rotavirus-antigen-positive samples from case children, and rotavirus qRT-PCR-positive samples from household members with a Ct value of ≤35, underwent G- and P-typing using a two-stage PCR with consensus and random primers^17^. Anti-rotavirus IgA geometric mean titres (IU/ml IgA) were measured using a semi-quantitative sandwich ELISA^18^ and were calculated using a minimum of two values per sample with a coefficient of variation (CV) <20%. Results were defined as zero if below the lower limit of detection. Clinical disease in household contacts was defined as any reported vomiting or diarrhea during the follow-up period.

### Statistical analysis

Distributions of continuous variables were examined and categorical variables were tabulated to generate descriptive statistics. Missing observations were assumed to be missing completely at random. Two-sided t-tests were used to compare independent means of normally distributed data, and rank-sum tests were used to compare non-normally distributed data. Chi-squared or Fischer’s exact tests were used to compare categorical variables.

Risk factors for rotavirus transmission within case households were identified using logistic regression models with a random effect to account for household-level clustering. A conceptual framework (Figure 1) was developed to account for the hierarchical relationship between predictive variables^19^. Variables were divided into two initial groups: those relating to the infectiousness of the symptomatic case child and those relating to the susceptibility of household contacts. Susceptibility variables were further divided into proximal susceptibility (individual level), and distal susceptibility (household level) variables. Individual models were built for each group to identify risk factors of importance whilst adjusting for potential confounding. A final model was then built incorporating all three groups, beginning with distal susceptibility variables then adding proximal susceptibility variables and infectiousness variables. Separate models were built for infection and disease. All variables with p<0·1 on univariable analysis were tested for inclusion in the final models. Nested models were compared using likelihood ratio tests. Variables were retained in the model if p<0·1 at any stage of the procedures outlined above.

**Figure 1.**
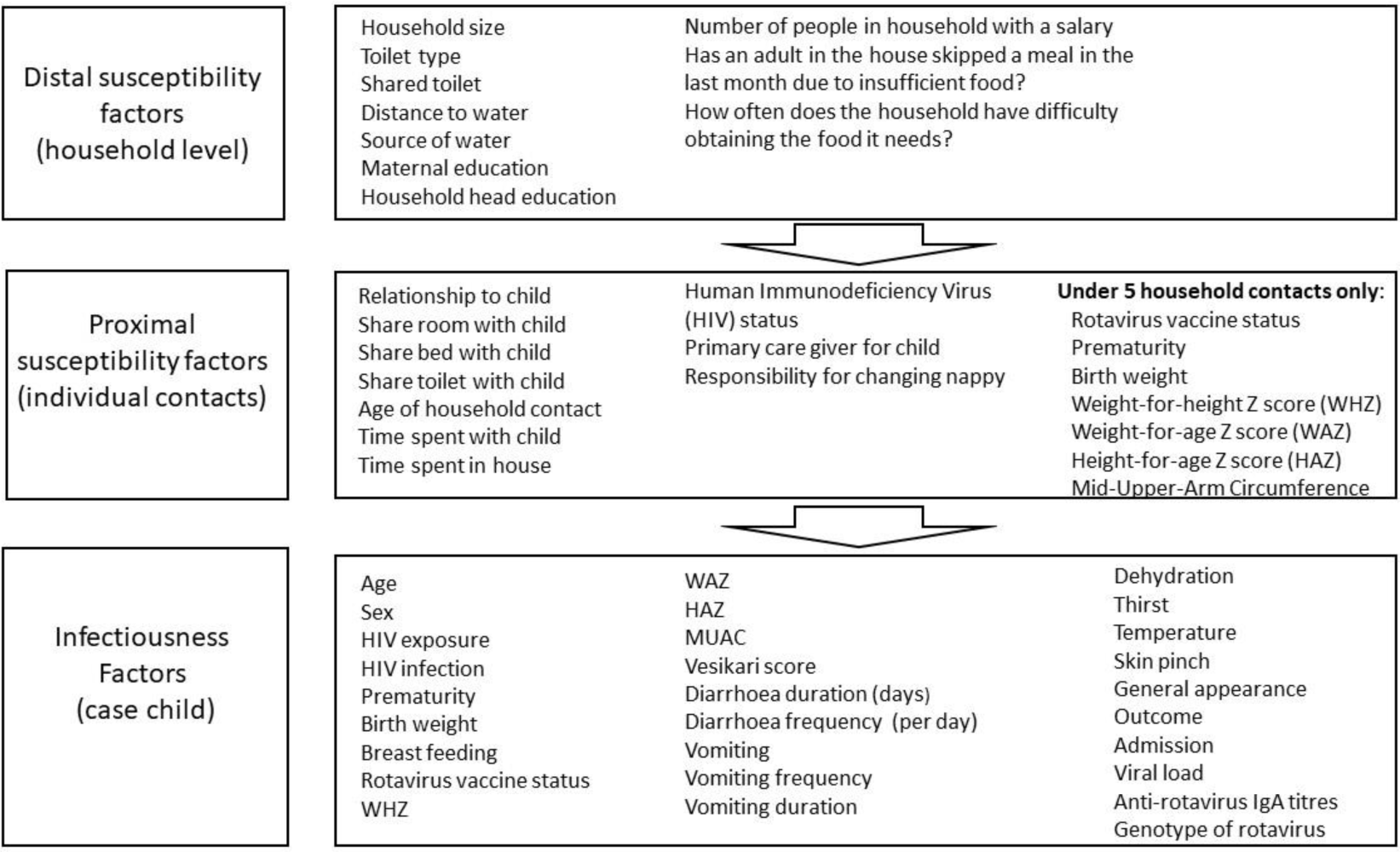
Conceptual framework for multivariate model.

## Estimating vaccine effectiveness against transmission (VE_T_)

Vaccine effectiveness against transmission of rotavirus infection (VE_T_) was estimated using the following approach:

1. Children were divided into three severity groups: very severe disease (Vesikari score ≥15), less severe disease (Vesikari score <15) and asymptomatic infection. SAR for infection among household contacts of index cases with very severe (*V*) and less severe (*M*) disease and of asymptomatic children (*A*) was estimated using data from the household transmission study. The prevalence of asymptomatic infection in household members of control households was used as an estimate of SAR for asymptomatic children, accepting that this is likely to be an over-estimate. Receiver operating characteristics (ROC) analysis together with Youden’s index^20^ was used verify that a cut-off of severity score 15 had good discriminatory power in differentiating risk of transmission (Figure S1).
2. We assumed that receipt of rotavirus vaccination would have resulted in children from the “very severe” disease category moving into the “less severe” category and children from the “less severe” category moving into an “asymptomatically infected” category at a proportion determined by the respective VE estimates (Figure 2). The number of vaccinated children with very severe (*SV*) and less severe (*MV*) disease was observed in the household transmission study. The number of children with very severe rotavirus disease in a hypothetical unvaccinated population (*SU*) was estimated as *SU*= *SV*/(1-*VE*_*S*_), where *VE*_*S*_ is the vaccine effectiveness against very severe disease. Similarly, the number of children with less severe disease in an unvaccinated population (*MU*) was estimated as *MU*= *MV*/(1-*VE*_*M*_), where *VE*_*M*_ is the vaccine effectiveness against less severe disease. The total number of children with very severe or less severe rotavirus disease in an unvaccinated population was therefore:

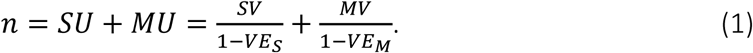
3. Vaccine effectiveness against very severe and less severe rotavirus disease were calculated as 1 minus the odds ratio (OR) of 2 versus 0 doses of vaccine. ORs were estimated using logistic regression models fit to data from the rotavirus surveillance platform in Blantyre, Malawi. Under idealised test-negative case-control designs, the OR provides an unbiased estimate of the relative risk (RR)^21^;
4. Vaccine effectiveness against transmission (*VE*_*T*_) was estimated as 1 minus the ratio of the SARs in vaccinated and unvaccinated populations using the following equation:

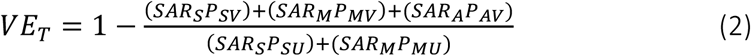

where *SAR*_*x*_ is the household secondary attack rate of infection for a household with an index child experiencing infection severity *x*, and *P*_*xy*_ indicates the proportion of index children (cases or controls) with rotavirus infection of severity *x* and vaccination status *y*. Proportions were estimated as: *P*_*SV*_ = *SV*/*n, P*_*MV*_ = *MV*/*n, P*_*AV*_=(*n*-*SV*-*MV*)/*n*), *P*_*SU*_ = *SU*/*n, P*_*MU*_ = *MU*/n.

Bootstrapped 95% confidence limits for *VE*_*T*_ were generated by sampling 10,000 times from the distributions of the corresponding parameters. Bootstrap samples of VE (1-OR) for very severe and less severe disease were generated by sampling from log-normal distributions using the mean and standard deviation derived from logistic regression models in step 3.

## Ethics

The study was approved by the University of Liverpool Research Ethics committee (# 000757), and the Malawi College of Medicine Research Ethics Committee (P.09/14/1623).

**Figure 2.**
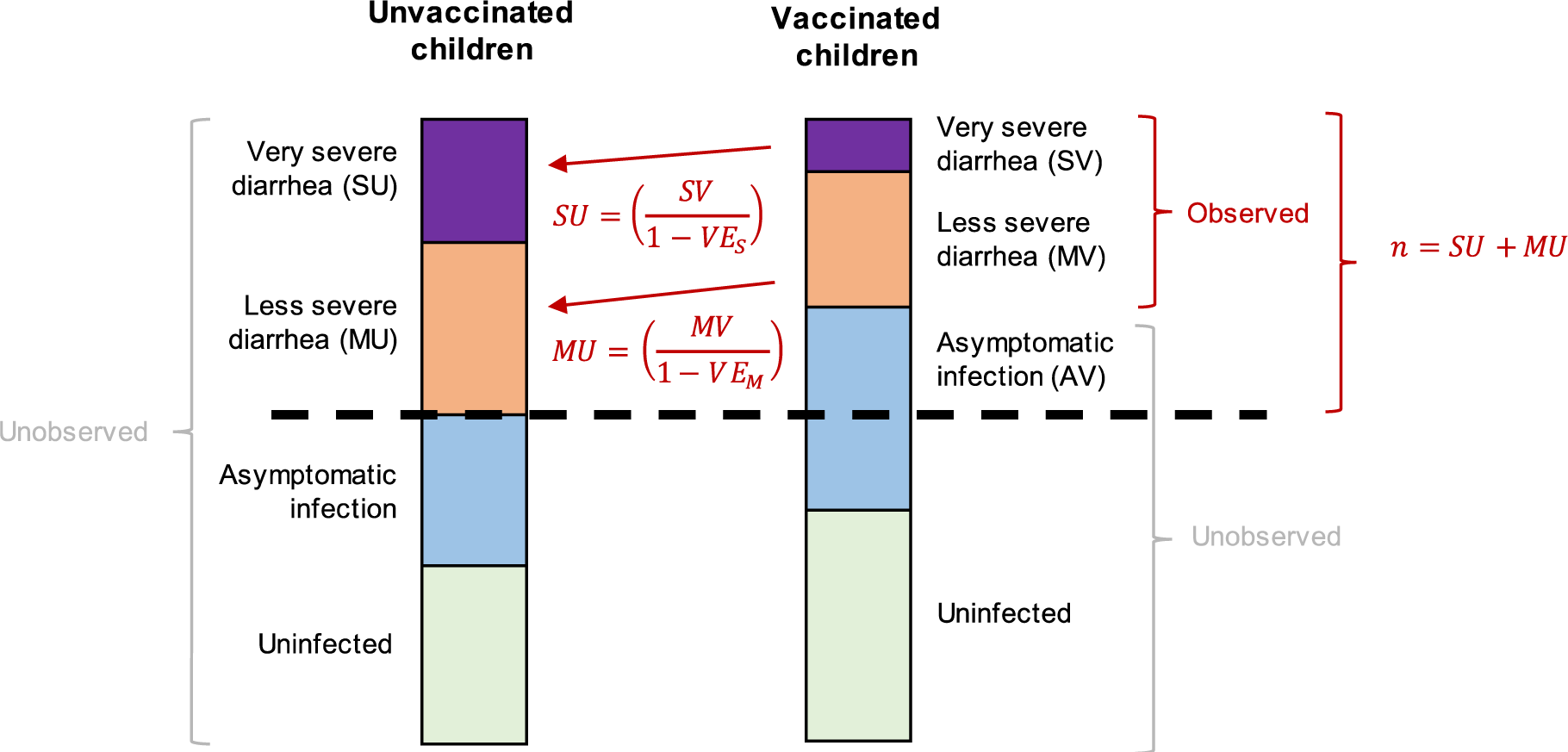
Conceptual diagram of the effect of vaccination on the proportion of children with rotavirus infection of different degrees of disease severity. In our study, we observed cases of very severe (*SV*) and less severe (*MV*) rotavirus diarrhoea among vaccinated children. We used the estimated vaccine effectiveness against very severe (*VE*_*S*_) and less severe rotavirus diarrhoea (*VE*_*M*_) to infer the number of very severe (*SU*) and less severe (*MU*) rotavirus cases in an unvaccinated population. The dashed line represents the size of the potentially observable population (*n*). Our analysis is conservative in that it assumes that the proportion of children who are asymptomatically infected or uninfected in a vaccinated population is greater than or equal to the corresponding proportions in an unvaccinated population.

## Results

### Characteristics of index children and households

A total of 196 case households containing a symptomatic rotavirus-positive case child, and 55 control households containing a frequency age-matched asymptomatic child were recruited. Median age was 11·5 months among both cases and age-matched control children; 55% of rotavirus-positive case children and 47% of control children were males (p=0.3) (Table 1). Rotavirus vaccine coverage was high (≥99.0% among both cases and controls). Household characteristics are listed in Tables S1 and S2. Anti-rotavirus IgA titres in case children at presentation were low (median 4 IU/ml). Viral loads in case children were high, with a median Ct value of 19.1 (interquartile range (IQR) 17.2, 22.2) corresponding to median copy numbers of 1.67 × 10^7^ (IQR 1.63 × 10^6^, 6.37 × 10^7^). Viral loads in household contacts were markedly lower; median Ct value was 34.8 (IQR 31.8, 36.6), corresponding to a median viral load of 712 (IQR 256, 3704).

**Table 1.**
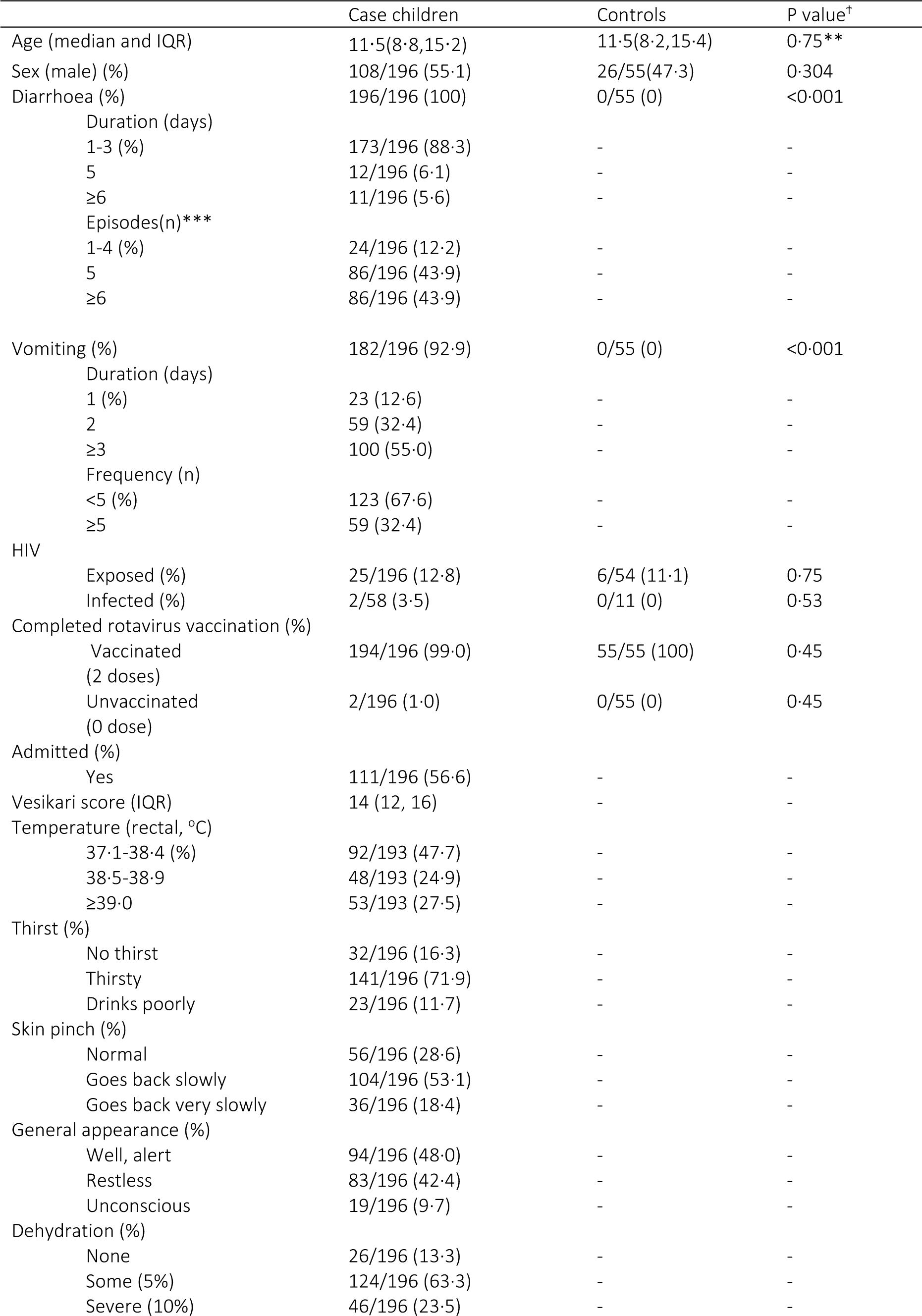
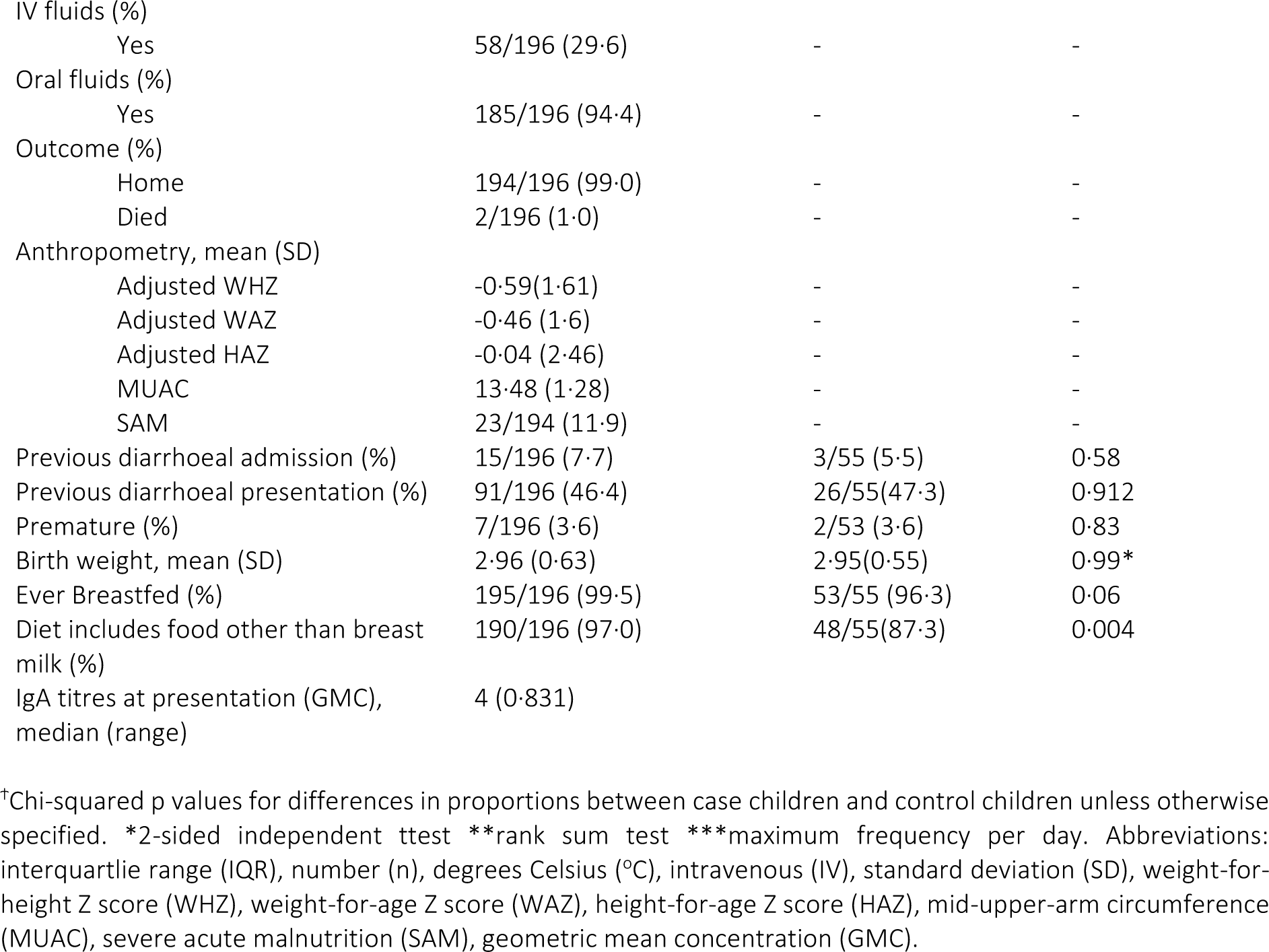
Characteristics of index and control children.

### Secondary attack rates

A total of 705 household members were recruited from 196 case households. At least one faecal sample was collected from 665 individuals from 188 case households, with a total of 1212 samples collected. The SAR for infection among household contacts of case children was high, with 65% of individuals positive for rotavirus (Table 2). Secondary attack rates for disease were much lower, with 48/699 (7%) household contacts reporting symptoms of gastroenteritis. Of these, 47 had samples available for testing and 37 (77.1%) were positive for rotavirus, resulting in a SAR for clinical rotavirus disease of 37/698 (5.3%). Rates of clinical disease were significantly higher among children <5 years (12/91, 13.2%, p<0.001).

**Table 2.** Secondary attack rates for rotavirus infection and clinical disease.

|  | Infection |  |  | Clinical rotavirus disease |  |  |
| --- | --- | --- | --- | --- | --- | --- |
|  | Case households | Control households |  | Case households | Control households | P-value* |
| Overall | 434/665 (65.3%) | 40/144 (27.8%) | <0.001<sup>†</sup> | 37/698 (5.3) | 0/153(0.0) | 0.004<sup>†</sup> |
| Age stratified (years) |  |  |  |  |  |  |
| 0-4 | 57/88 (64.8) | 2/10 (20.0) | 0.006<sup>†</sup> | 12/91 (13.2) | 0/11 (0.0) | 0.200<sup>†</sup> |
| 5-14 | 127/193 (65.8) | 14/48 (29.2) | <0.001<sup>†</sup> | 4/197 (2.0) | 0/53 (0.0) | 0.296<sup>†</sup> |
| 15-45 | 240/367 (65.4) | 5/20 (27.4) | <0.001<sup>†</sup> | 20/390 (5.1) | 0/87 (0.0) | 0.031<sup>†</sup> |
| 45+ | 9/16 (56.3) | 1/2 (50.0) | 0.867<sup>†</sup> | 1/18 (5.6) | 0/2 (0.0) | 0.732<sup>†</sup> |
| χ<sup>2</sup> p* | 0.894 | 0.838 |  | 0.001 | NA |  |

\*Chi-squared p value for difference in proportion across age categories
<sup>†</sup>Chi-squared p value comparing difference in proportion between cases and controls

The prevalence of rotavirus shedding in the control households was 27.8% (40/144), significantly lower than in case households (Table 2).

### Rotavirus genotypes

Of 195 samples from case children with available genotyping data, almost a third were genotype G2P[4] (30.8%). The next most frequent genotypes were G1P[8] (24.6%), G2[P6] (14.4%) and G12P[6] (8.2%) (Figure S2). A total of 297 samples were genotyped from household contacts of cases; in almost one third (94/297, 31.6%) both G and P types were the same as those identified in the case child. Comparison of genotypes in case children and household contacts are reported in Table S3. Only seven samples from control households could be genotyped due to low viral load in most control samples (data not shown).

### Risk factors for transmission of rotavirus infection

Increasing disease severity (per Vesikari score unit) in the case child was strongly associated with transmission to household contacts (OR 1.17, 95% CI 1.06, 1.30; p=0.003, Table 3). MUAC in the case child was also positively associated with risk of transmission. At the household level (distal susceptibility factors), having at least one household member with a regular salary was associated with reduced susceptibility to infection. Conversely, a recent history of difficulty obtaining sufficient food for the household was also associated with reduced susceptibility to infection. At the individual level (proximal susceptibility factors), there was strong evidence that the proximity of relationship with the case child was associated with risk of transmission, with mothers significantly more likely to become infected with rotavirus than other adult relatives or child household contacts (Table 3). Univariate associations for risk of transmission of infection are detailed in Tables S4-S6.

**Table 3.** Risk factors for transmission of rotavirus infection.

| Variable | OR* | P value | 95% CI |
| --- | --- | --- | --- |
| Infectiousness risk factors |  |  |  |
| Vesikari score in index child | 1.17 | 0.003 | 1.06, 1.30 |
| MUAC* in index child | 1.32 | 0.015 | 1.06, 1.66 |
| Distal susceptibility factors |  |  |  |
| Number of adults with salary in household |  |  |  |
| None | REF |  |  |
| ≥1 | 0.44 | 0.007 | 0.24, 0.80 |
| Problems getting food in the past month (%) |  |  |  |
| No | REF |  |  |
| Sometimes/often | 0.58 | 0.08 | 0.31, 1.06 |
| Proximal susceptibility factors |  |  |  |
| Relationship with child |  |  |  |
| Mother | REF |  |  |
| Other adult relative | 0.29 | 0.000 | 0.16, 0.50 |
| Child contact | 0.44 | 0.002 | 0.26, 0.74 |

\*OR indicates odds ratio, MUAC indicates Mid Upper Arm Circumference. Odds ratio for Vesikari score and MUAC are per variable unit

### Risk factors for transmission of rotavirus disease

The primary infectiousness risk factor was disease severity in the index case, with a positive association between increasing disease severity and risk of clinical rotavirus disease in household contacts (Table 4). There was evidence of a weak association between genotype of rotavirus in the index child and risk of disease transmission, with an increased risk of transmission with G1P[8] genotype compared to G2P[4], G2P[6] and G12P[6]. At the household level (distal susceptibility factors), use of a pit or water toilet was associated with a reduced odds of rotavirus disease compared to having no toilet, though this association was no longer statistically significant in the final model. At the individual level (proximal susceptibility factors), age of the household contact was significantly associated with risk of disease; children aged <5 years had the greatest risk of disease compared to children aged 5-15 years and older adults. Univariate associations for risk of disease are presented in Tables S7-S9.

**Table 4.** Risk factors for rotavirus disease transmission.

|  | OR | P value | 95% CI |
| --- | --- | --- | --- |
| Infectiousness risk factors |  |  |  |
| Vesikari score in index child | 1.28 | 0.005 | 1.08, 1.52 |
| Rotavirus genotype, index child |  |  |  |
| G1P[8] | REF |  |  |
| G2P[4] | 0.36 | 0.059 | 0.12, 1.03 |
| G2P[6] | 0.43 | 0.219 | 0.11, 1.64 |
| G12P[6] | 0.82 | 0.759 | 0.23, 2.95 |
| Other | 1.10 | 0.842 | 0.44, 2.71 |
| Distal susceptibility factors |  |  |  |
| Toilet type |  |  |  |
| None | REF |  |  |
| Simple pit/VIP* | 0.56 | 0.484 | 0.11, 2.83 |
| Water toilet | 0.42 | 0.520 | 0.03, 5.96 |
| Proximal susceptibility factors |  |  |  |
| Household member age |  |  |  |
| <5 years | REF |  |  |
| 5-15 years | 0.13 | 0.001 | 0.04, 0.46 |
| 15-45 years | 0.38 | 0.031 | 0.17, 0.92 |
| 45+ years | 0.35 | 0.351 | 0.04, 3.22 |

\*VIP= Ventilated improved pit latrine, OR indicates odds ratio. Odds ratio for Vesikari score is per variable unit

### Vaccine Effectiveness against Transmission (VE_T_)

Vaccine effectiveness against very severe and less severe rotavirus disease were estimated to be 69% (95% CI: -10, 91%) and 56% (95% CI 3, 79%), respectively, based on the diarrhoeal surveillance dataset. As a result, we estimated that the proportion of unvaccinated children with very severe disease would be 0.44 (95% CI 0.15, 0.78) and the proportion of unvaccinated children with less severe disease would be 0.55 (95% CI 0.22, 0.85). In comparison, the proportion of vaccinated children with very severe and less severe disease were estimated to be 0.13 (95% CI 0.06, 0.27) and 0.23 (95% CI 0.11, 0.47), respectively, and the proportion of vaccinated children who became asymptomatic was estimated to be 0.63 (95% CI 0.27, 0.83). The SARs for severe disease, less severe disease, and asymptomatic infection were estimated to be 72% (95% CI 64%, 79%), 64% (95% CI 57%, 69%), and 25% (95% CI 16%, 35%), respectively. Combining this information in equation 2, we estimated a VE_T_ of 39% (95% CI 16%, 57%) (Figure 3).

**Figure 3.**
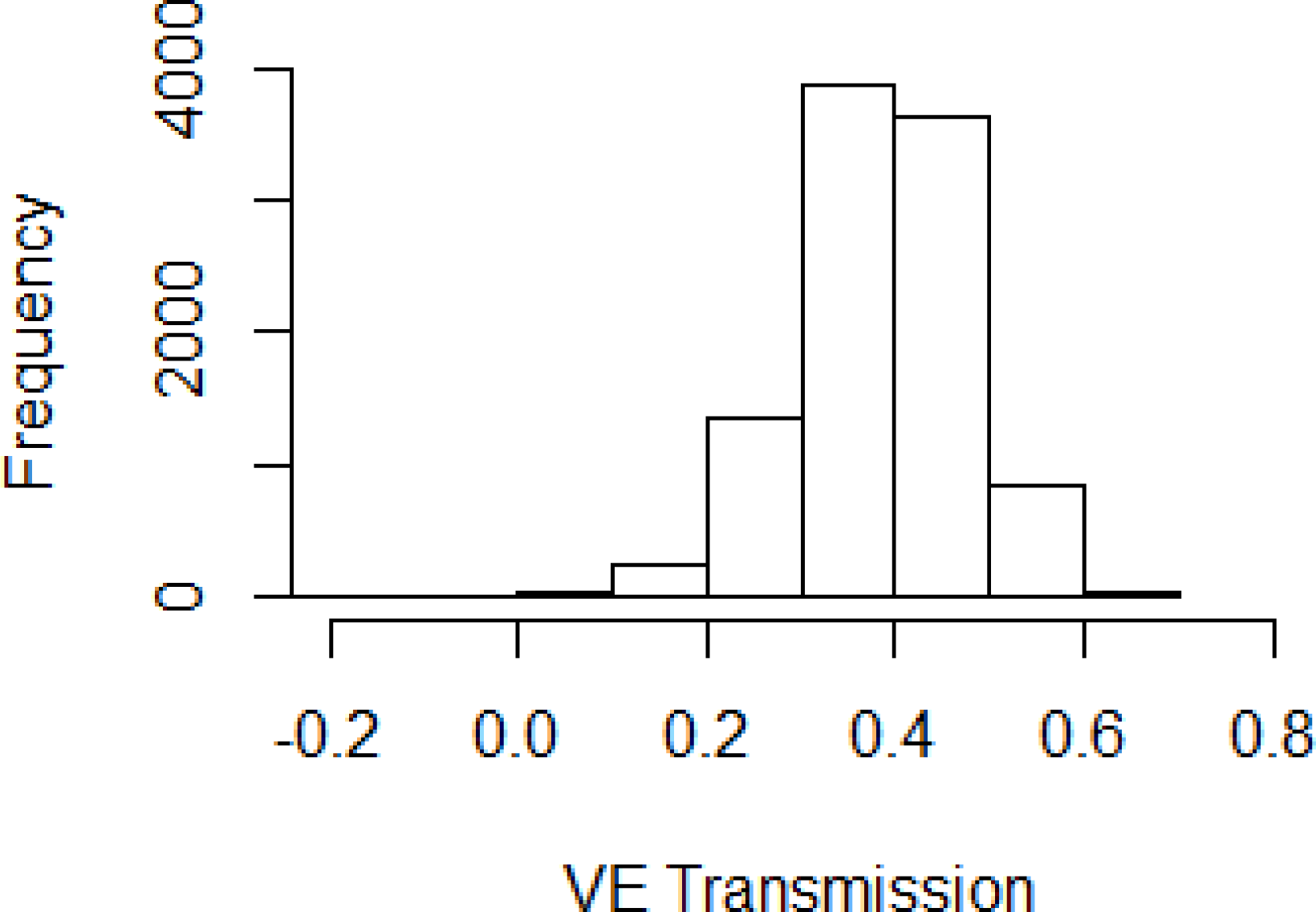
Distribution of estimates for vaccine effectiveness against transmission (*VE*_*T*_). The histogram of 10,000 bootstrap samples of the estimated *VE*_*T*_ is plotted.

## Discussion

In Malawi, very high attack rates (65%) for rotavirus infection were observed in households following contact with a symptomatic rotavirus case despite very high rotavirus vaccine coverage. However, the attack rate for rotavirus disease was much lower (5%). Disease severity in the index child was an important predictor of transmission of both infection and disease to household contacts. We estimated a VE against transmission of rotavirus infection (VE_T_) of 39%.

The majority of case children in our study represent rotavirus vaccine failures; high viral shedding density and low anti-rotavirus IgA titres at the time of presentation likely explains the high SAR. Our estimate of SAR for rotavirus infection is consistent with studies from New Zealand and Ecuador, which reported SARs for rotavirus infection of 48% and 55%, respectively^9,10^. In contrast, we observed much lower overall attack rates for clinical disease (5%), compared with 15% and 38% reported for Ecuador and New Zealand, respectively. This difference could be explained by a high background force of rotavirus infection in Malawi resulting in frequent “boosting” of immunity against clinical disease, particularly among older children and adult; of note, the clinical SAR was highest in children aged 0-4 years^22^. We also observed a high frequency of rotavirus infection (28%) in control households. Whilst this is substantially greater than observed in higher income settings, such as the UK and Ecuador^9,23^, it is consistent with published studies from sub-Saharan Africa^24,25^, and is plausible given the high force of rotavirus infection in Malawi ^22^and high levels of poverty, crowding, and poor access to water and sanitation. The inconsistencies between rotavirus genotypes detected in case children and their household contacts may also reflect the high frequency of asymptomatic shedding identified in the community^9^.

Increasing disease severity was associated with increased odds of rotavirus transmission for both infection and disease; by reducing disease severity, rotavirus vaccine has the potential to reduce the infectiousness of a symptomatic index case even in the event of clinical vaccine failure. This is a phenomenon described with other pathogens such as pertussis, but not as yet with rotavirus^26^. In this study, we estimated that in a semi-urban population in Malawi, with extremely high rates of rotavirus transmission, rotavirus vaccine reduces population-level rotavirus transmission by 39%. Such a reduction in transmission has the potential to make a considerable impact on the burden of rotavirus disease in the community, and is consistent with previous estimates of indirect effectiveness of rotavirus vaccination from Malawi from hospital-based studies and mathematical models^22,27^. In contrast, horizontal transmission of vaccine virus is unlikely to make a significant contribution to indirect effects in this setting, since vaccine transmission within households is rare^28^.

Lack of a regular salary in the household increased the risk of transmission of infection. This likely reflects relative poverty, which could increase rotavirus transmission for reasons including crowding, sanitation, or carer education levels, or other unmeasured factors. Close contact and proximity of relationship to the index child also increased the risk of transmission of infection, emphasising the role that personal/hand hygiene and behavioural measures may play in preventing rotavirus transmission within households. Rotavirus genotype G1P[8] was weakly associated with an increased risk of transmission, consistent with the global predominance of G1P[8] prior to widespread introduction of vaccination.

Our study has important limitations. Firstly, the direction of infection cannot be defined with certainty using this study design, since households were recruited only when an index child presented with rotavirus gastroenteritis; however, the pattern of transmission supports young infants bringing rotavirus into the household. Secondly, our estimate of *VE*_*T*_ assumes that asymptomatic infections contribute a relatively small amount to the overall SAR, and that the prevalence of asymptomatic infection is similar or lower in an unvaccinated population. The prevalence of asymptomatic infections among vaccinated infants and VE against asymptomatic infection are unknown. Furthermore, *VE*_*T*_ is dependent on locally-specific parameters, and our estimate may not be generalisable to other settings.

Despite sustained high coverage of rotavirus vaccine in Malawi and other low-income African countries, the burden of rotavirus disease remains high. We demonstrated frequent transmission of rotavirus infection from symptomatic, vaccinated children in Malawi to their household contacts and high background rates of asymptomatic rotavirus shedding. Our estimate of rotavirus vaccine effectiveness against transmission of infection of 39% suggests indirect protection has the potential to substantially contribute to vaccine impact on community disease burden. Together with direct vaccine effectiveness estimates, indirect or “herd” protection should be considered in future health economic assessments of rotavirus vaccine impact. This may be particularly relevant in low-income, high-disease-burden environments.

## Data Availability

All data used in manuscript will be made publically available via university data repository

## Contributors

AB had overall responsibilty for study design and execution, data analysis, and manuscript drafting, editing and writing. LP and NBZ contributed to study design, data collection and manuscript editing and writing. KCJ contributed to laboratory data collection and supervision and manucript editing. BL contributed to study design and data analysis. MIG contributed to study design, laboratory data collection and supervision, and manuscript editing. VEP contributed to study design, data analysis and manuscript editing and writing. NAC contributed to study design, data collection, and manuscript editing and writing.

## Acknowledgements

This study was supported by two Wellcome Trust Clinical PhD Fellowships [grant numbers 102466/Z/13/A to AB and 102464/Z/13/A to LP], a Wellcome Trust Programme Grant [grant number 091909/Z/10/Z], the MLW Programme Core Grant Strategic Award [grant number 101113/Z/13/Z] and by the U.S. National Institutes of Health/National Institute of Allergy and Infectious Diseases [grant number R01-AI112970 to VEP]. K.C.J. is supported by an International Wellcome Trust Training Fellowship (grant number: 201945/Z/16/Z). We thank all infants and their families who participated and all members of the RotaRITE study team. We are grateful for the support of the Malawi Ministry of Health and clinical staff at the recruitment sites.

